# Aging in India: Comparison of Conventional and Prospective Measures, 2011

**DOI:** 10.1101/2022.04.11.22273700

**Authors:** Ankita Srivastava, Saikia Nandita

## Abstract

Conventional measurements of aging do not take into account the dynamic nature of aging-related characteristics over time. Therefore, in order to refine the estimates of aging, demographers have proposed prospective measures based on remaining life expectancy, Sanderson and Scherbov (2007). We compared these new measures with conventional aging measures using the data from the Census of India 2011 and Sample Registration System life tables 2009–2013. In conventional aging measures, we used life expectancy at age 60 and the old age dependency ratio (OADR), whereas for new measures of aging, we applied the threshold of old age based on the remaining life expectancy and prospective old age dependency ratio (POADR). Both measures of aging provided different estimates of the aging population at the national and subnational levels. At national level, application of prospective measures increased the number of older dependents from 66.4 million to 71.8 million (OADR: 8.6% vs. POADR: 10.6%). We observed profound variation at sub-national level in India. We also observed that the prospective ageing measures not only provided higher estimates of ageing burden in India, but also altered the gender and rural urban differential in ageing. Considering the heterogeneity of life expectancies across Indian states, prospective measures provide more accurate refined estimates of aging burden in India as they are based on length of life expectancy. Application of these measures has great policy relevance in India.

## Introduction

The total population aged 60 and above is growing much faster in developing regions than in developed regions. Application of standard measurements on ageing employed by the United Nations (UN) shows that developing regions are home to a growing share of the world’s older population (United Nations 2017a). For the next 50 years, the number of individuals aged 65 and over is predicted to surge across Asia (United Nations 2017b). The number of people aged 60 and above in India is currently 549 million and is anticipated to rise to 1.3 billion by 2050 (United Nations 2017b). India, one of the world’s two population superpowers, will soon face the ageing burden. In recent decades, India’s lifespan has increased while its fertility has decreased, resulting in 69 million elderlies. Moreover, according to the most recent UN statistics, the share of the population aged 60 and over is projected to increase from 8% to about 20% by 2050. They also claimed that by 2050, the elderly will outnumber children under age 15 (UNDESA 2019) and constitute 34% of the total population (UNFPA 2017). These results corroborate several earlier research (e.g., James, 1994). The standard old age dependency ratio (OADR) increased from 10.9 percent in 1961 to 14.2 percent in 2011, and it varies greatly by location (rural vs. urban) and geographical region.

To better understand the ageing process, several studies have evaluated the concept of healthy ageing, which encompasses a wide spectrum of diseases (Balachandran and James 2019; Balachandran *et al*. 2019; Chang *et al*. 2019). The existing literature takes two approaches to ageing measurement: the first set focuses on changes in age structure without regard for morbidity (d’Albis and Collard 2013); this includes metrics such as age structure shifts to older ages (Gavrilov and Heuveline 2003), increases in population median age (United States Census Bureau 2017), increases in life expectancy, remaining years to live (Ryder 1975; Gavrilov and Heuveline 2003), and changes in old age dependency ratios (UNDESA 2017a). The second group is concerned with assessing the functional state of the elderly population by objective measurements such as frailty (Fried *et al*. 2001; Mitnitski, Mogilner and Rockwood 2001; Searle *et al*. 2008), biomarkers (Belsky *et al*. 2015), and cognitive functioning (Skirbekk, Loichinger and Weber 2012), as well as subjective indicators such as self-reported health and instrumental restriction of activities of daily life. The second approach is outside the scope of this study, since the sole purpose of this article is to assess the elderly dependence situation in various states of India via the lens of demographics and changes in life expectancy.

Therefore, continuous research and debate on the consequences of ageing in India are ongoing. The massive challenge due to the growing burdens of noncommunicable diseases and disability, vulnerabilities of the female elderly, and insecurity about income have been intensively discussed in ongoing research in India (Syam Prasad 2011; Prakasam 2012; Agarwal *et al*. 2016; Saikia *et al*. 2016; Arokiasamy 2018; Parmar and Saikia 2018).

Bhagat and Unisa described an improvement of the dependency measurement. They employed three types of dependency: old age economic, adult, and relative. Using these dependency ratios, they investigated inter-state and gender variances. The findings demonstrated that geriatric relative dependence is lower than adult relative dependence, and the potential to harvest the demographic window in India is being squandered Bhagat and Unisa (2006).

According to Irudaya et.al (1996), the standard dependency ratio estimate based on the 60+ non-working population should be revised to determine the precise level of dependent burden by only considering non-workers above the age of 60, and revising the retirement age. According to Visaria, (2001), it is difficult to estimate the number of elderly individuals and their actual needs. The study emphasized health care demands and available resources, as well as financial instability among the elderly, which was more pronounced in females than males. Moreover, financial issues usually affect widows and elderly nuclear families (James 1994; Meijer 2012; Kalita 2017).

Subaiya and Bansod (2011) also presented essential facts and patterns about historical and future population ageing in India utilizing median age, ageing index, and OADRs. They discovered that India’s age structure is changing from young to old. Their study’s index of ageing was expressed as the number of people over 60 per 100 children below 15. In 2001, the index of ageing was 23.4 elderly per 100 children, but this figure is projected to 53 elderlies per 100 children by 2026, indicating an accelerating ageing trend. The authors also highlighted rising OADRs, falling potential support ratios, and feminization of ageing in India.

There have also been attempts to address geographical differences in ageing in India (Das, Sengupta and Paul 2018), and challenges in access and affordability of health among the elderly (Dey et al., 2012). The considerable interstate discrepancy in ageing in India is highlighted in the UNFPA 2017 study. All southern states have a higher elderly population, in addition to a few northern states such as Himachal Pradesh, Maharashtra, Odisha, and Punjab. According to the 2011 Census, these states have a higher percentage of people aged 60 and above than the central and northern states (UNFPA 2017). However, according to Bhat (2004), the problem of the elderly (65+) is not state-specific, as the elderly population is increasing rapidly even in northern states.

However, demographers have argued that the conventional measurement of ageing is a static concept that ignores human characteristic improvement with time. Modern diet, medical technology, and lifestyle advancements allow a 70-year-old to function or behave like a 60-year-old. Sanderson and Scherbov (2007) presented a new forward-looking concept of age, called, “prospective age”. Rather than chronological age, they developed characteristic-equivalent ages, which are defined as the age at which a measure of certain qualities remains constant. While conventional age is defined as “the number of years an individual has lived since his date of birth,” prospective age is defined as “the number of years a person is expected to live.” Hence, prospective age is the age at which everyone in a population has the same number of years left to live. These new population ageing measures are useful due to improved health and life expectancy.

Despite the increasing popularity of this new concept of ageing (Basten, 2013; Ediev, Sanderson, & Scherbov, 2019; Gietel-Basten, Scherbov, & Sanderson, 2015; Lutz, 2016; Sanderson & Scherbov, 2005; Sanderson & Scherbov, 2010; Sanderson, Scherbov, & Gerland, 2017; Sanderson & Scherbov, 2008; Scherbov, Sanderson, & Samir KC, 2014; Scherbov & Sanderson, 2016; UNDESA, 2017), to the best of our knowledge, it has not been discussed in the context of India, which is known for unusual diversity in demographic characteristics.

Our literature review reveals that research on ageing in India have estimated the absolute and relative burden of ageing using either the percentage ageing (60 or 65+) or the standard OADR. Generally, researchers in this field have focused on negative implications of ageing, such as low labor participation and excessive social expenditure (Liebig and Rajan 2003; Bloom, Canning and Fink 2010; Wolf *et al*. 2011; Dey *et al*. 2012). However, the elderly may be frail and incapable of physical labour, that does not mean they are unproductive.

Elders exhibit competence and expertise, as well as an emotional force capable of uniting and commanding discipline and respect. With proactive legislation, efforts, and a focus on the inherent characteristics of the elderly, ageing may be considerably more elegant.

World-wide initiatives like the Madrid International Plan of Action on Ageing (2002), the United Nations Proclamation on Ageing (2002), the Shanghai Plan of Action (2002), and the Macau outcome document (2007) have all promoted “Graceful Aging.” Achieving optimal physical, social, and mental health is the goal of graceful ageing (Mathuranath 2005). The WHO has termed this “Active Aging” since the late 1990s. The transition from a needs-based strategy to a rights-based participatory approach respects the elder population’s equal opportunity and treatment in all spheres of life (Chakraborti 2004).

Therefore, the aim of this study was two-fold. First, we estimated and compare the burden of ageing by applying the conventional and new measures of ageing by gender and type of residence, and then we examine the regional variation in ageing using these concepts.

## Data Source and Methodology

### Data Source

To achieve the paper’s objectives, we relied on data from two sources. We estimated the life expectancies of Indian states using SRS life table data from 2009–2013. The SRS was implemented as a pilot scheme in some Indian states in 1964–1965 to generate reliable national and state estimates of fertility and mortality. It was converted into a full-scale system in 1969–1970 and is the most reliable source of data on vital events in India and its major states (Bhat 2002; Saikia *et al*. 2011). We restricted our analysis to bigger states of India, for which SRS life tables are available.

The population data was collected from the 2011 Census of India, the most reliable source of demographic data since 1872. An extended de facto canvasser method is used in India for its decennial censuses. Individuals gather data through door-to-door visits for three weeks, and then collect data again to update the reference date and time. More details and data are available at the website of the Census of India (ORGI 2019).

### Methodology

A comparison of two ageing metrics was made. For conventional ageing measures, we used life expectancy at 60 and the standard OADR, whereas for prospective ageing measures, we used RLE and the prospective old age dependency ratio (POADR). Ryder (1975), defines age as the number of years left to live, not the number of years lived since birth. That study suggested that someone is “old” if their RLE is fewer than ten years. This sort of characterization distinguishes between individual and population ageing and may be used to define the proportion of older people in a population. Sanderson and Scherbov (2005, 2010) advocated standardizing the median age of the population for the expected remaining years of life. Instead of using age 60 or 65 as the threshold for old age, Sanderson and Scherbov (2005, 2010), propose a new threshold defined on the basis of RLE. The age at which average RLE equals 15 varies per demographic subgroup. They call this age the prospective age, and the measures that use them prospective measures of population ageing.

To compute the POADR by region and sex, we utilized population data from the 2011 Census of India and SRS abridged life tables for 2009–2013. Due to the uncertain quality of SRS mortality data beyond the age of 60 (Saikia *et al*. 2011), we used the Heligman-Pollard model to reconstruct age-specific mortality rates and life expectancy (Heligman and Pollard 1980). The old age threshold used in the computation of the prospective proportion of the population is age when RLE equal to 15.

Symbolically, it can be expressed as:

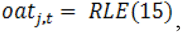

where oat_j,t_ is the old age threshold in state *j* in year *t*, and RLE (15) is the age in the life table for state j in year *t* where the RLE is equal to 15 years.

The conventional OADR was computed at the midpoint of 5-year intervals as follows:

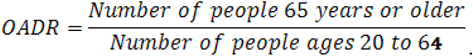

The POADR was calculated as follows:

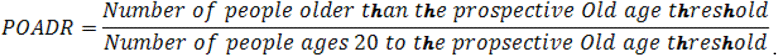

We computed both the conventional OADR and the POADR based on the Census of India 2011 data.

To compute conventional number of dependents in absolute terms, we multiplied the OADR by the working population aged 15–64, whereas, for prospective absolute number of dependents, we multiplied the POADR by the working population aged 15 to the prospective threshold age. Ageing was classified as high, moderate, or low in states where it was above the mean plus standard deviation (SD), between the mean and SD, or below the mean minus SD respectively.

## Results

### Regional variation in average remaining life years among various Indian states

As per **Figures 1 to 3** and **Table A1** in the appendix, the RLE (equals to 15 years) varies greatly amongst Indian states. The vertical bar shows the age at which people in each state have an RLE of 15. In Table 1, the value of 66.9 for Punjab implies that males in Punjab on average survive 15 years or less after reaching the age of 67.

**Figure 1.**
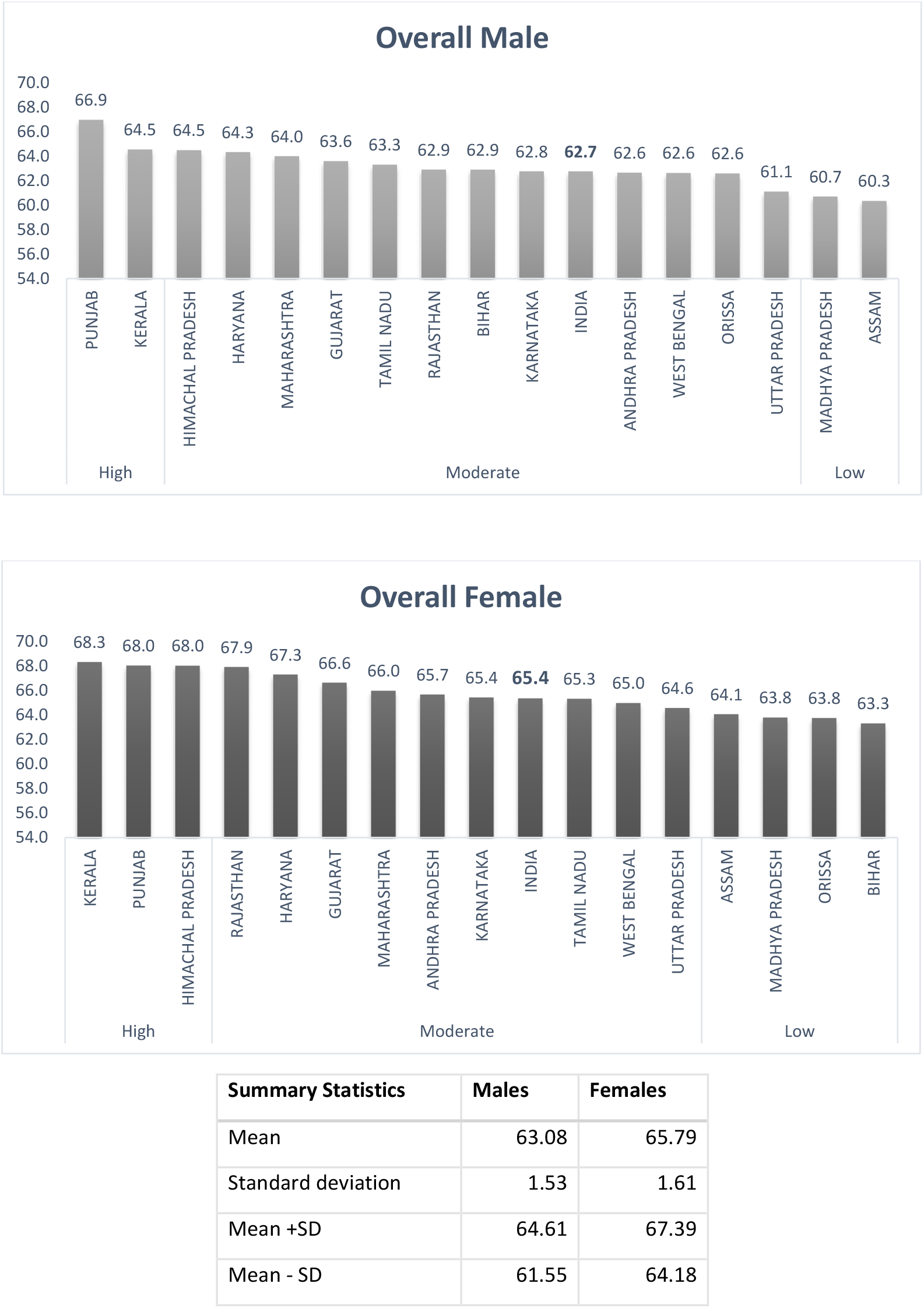
Regional variation in age when RLE=15 among various Indian states, total population, by sex, 2011. **Data source**: Calculation based on Sample Registration System Abridged Life Tables, 2009-2013

**Figure 2.**
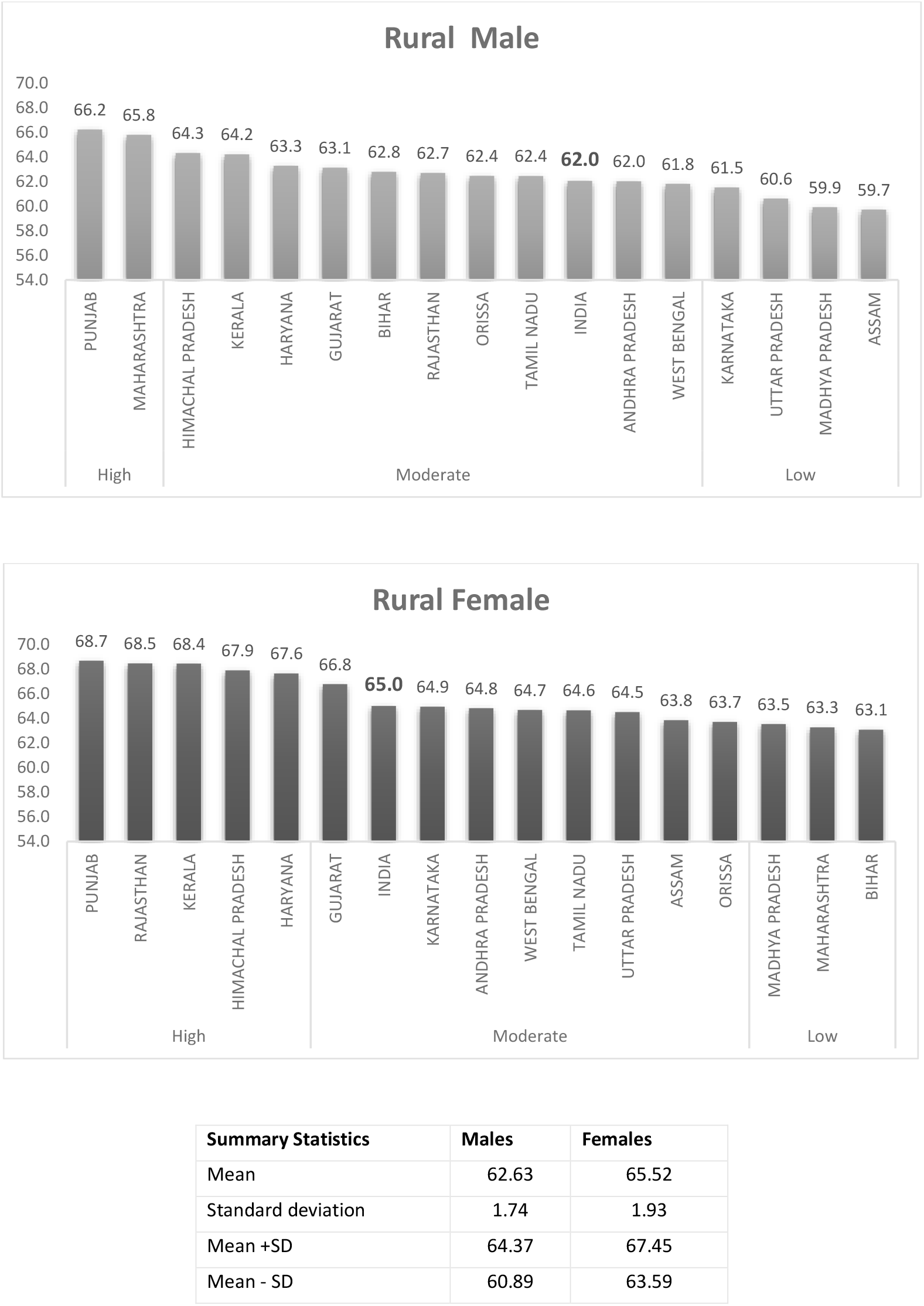
Regional variation in age when RLE=15 in the rural population of various Indian states by sex, 2011. **Data source**: Calculation based on Sample Registration System Abridged Life Tables, 2009–2013

**Figure 3.**
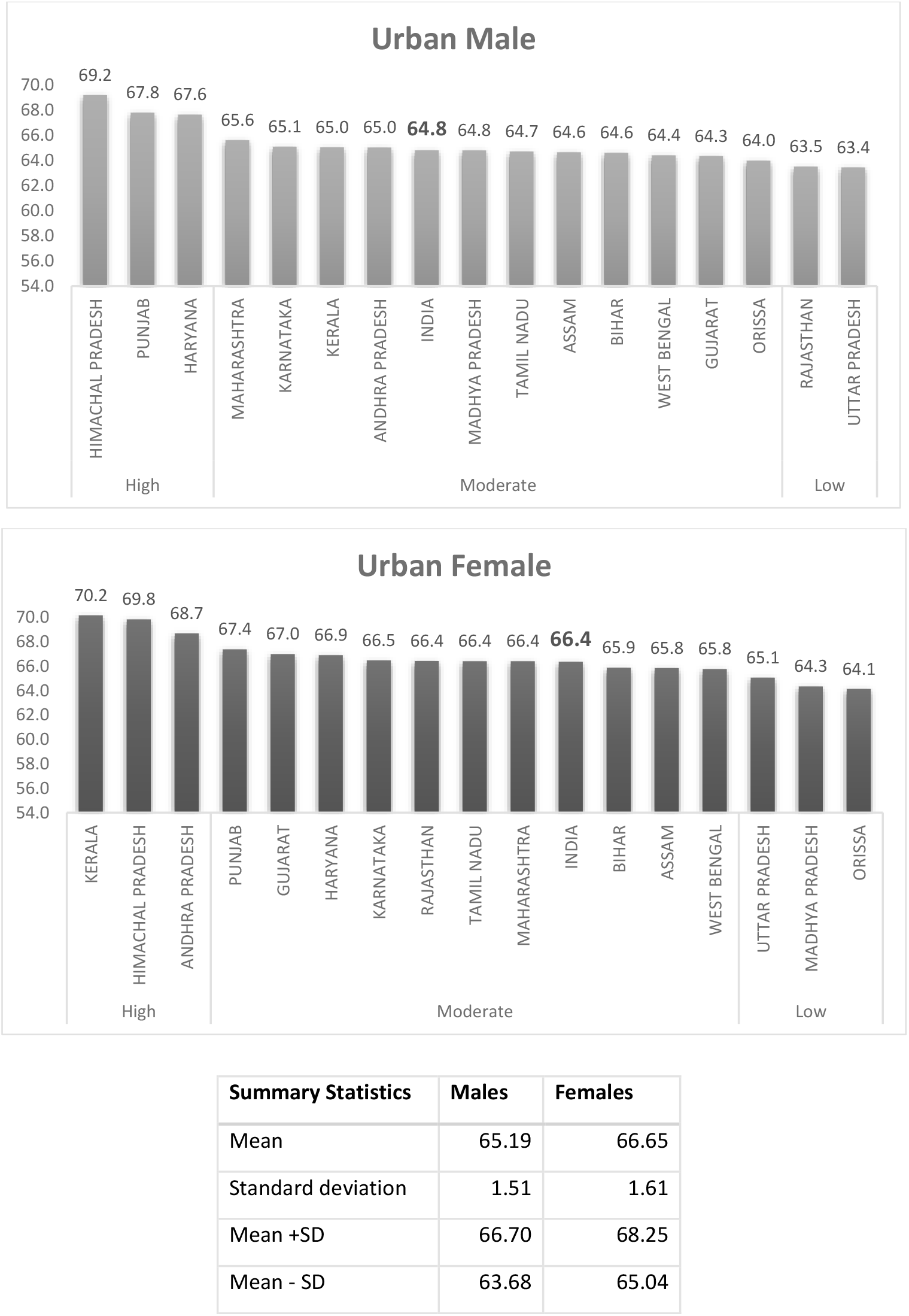
Regional variation in age when RLE=15 in the urban population of various Indian states by sex, 2011. **Data source**: Calculation based on Sample Registration System Abridged Life Tables, 2009–2013

**Table 1:** Comparison of state ranks based on LE60 and RLE15, by sex for the period 2009–2013.

| Male |  |  |  |  |  | Female |  |  |  |  |  |
| --- | --- | --- | --- | --- | --- | --- | --- | --- | --- | --- | --- |
| Level of ageing | States | RLE15 | New Ranks | LE60 | Old Ranks | Level of ageing | States | RLE15 | New Ranks | LE60 | Old Rank |
| High {> (Mean + Standard deviation)} | Punjab | 66.9 | 1 | 22.9 | 1 | High {> (Mean + Standard deviation)} | Kerala | 68.3 | 1 | 26.0 | 1 |
|  | Kerala | 64.5 | 2 | 21.8 | 3 |  | Punjab | 68.0 | 2 | 25.1 | 4 |
| Moderate (between Mean ± Standard Deviation) | Himachal Pradesh | 64.5 | 3 | 22.0 | 2 | | Himachal Pradesh | 68.0 | 3 | 25.2 | 3 |
|  | Haryana | 64.3 | 4 | 21.1 | 5 |  | Rajasthan | 67.9 | 4 | 25.3 | 2 |
| | Maharashtra | 64.0 | 5 | 21.6 | 4 | Moderate (between Mean ± Standard Deviation) | Haryana | 67.3 | 5 | 24.6 | 5 |
|  | Gujarat | 63.6 | 6 | 20.7 | 6 |  | Gujarat | 66.6 | 6 | 23.9 | 6 |
|  | Tamil Nadu | 63.3 | 7 | 20.7 | 6 |  | Maharashtra | 66.0 | 7 | 23.7 | 7 |
|  | Rajasthan | 62.9 | 8 | 20.6 | 9 |  | Andhra Pradesh | 65.7 | 8 | 23.1 | 8 |
|  | Bihar | 62.9 | 9 | 20.7 | 6 |  | Karnataka | 65.4 | 9 | 22.9 | 9 |
|  | Karnataka | 62.8 | 10 | 20.2 | 12 |  | Tamil Nadu | 65.3 | 10 | 22.9 | 9 |
|  | Andhra Pradesh | 62.6 | 11 | 20.2 | 12 |  | West Bengal | 65.0 | 11 | 22.8 | 11 |
|  | West Bengal | 62.6 | 12 | 20.6 | 9 |  | Uttar Pradesh | 64.6 | 12 | 21.9 | 12 |
|  | Odisha | 62.6 | 13 | 20.3 | 11 |  | Assam | 64.1 | 13 | 21.5 | 15 |
| Low {< (Mean - Standard deviation)} | Uttar Pradesh | 61.1 | 14 | 19.0 | 14 | Low {< (Mean - Standard deviation)} | Madhya Pradesh | 63.8 | 14 | 21.7 | 13 |
|  | Madhya Pradesh | 60.7 | 15 | 19.0 | 14 |  | Odisha | 63.8 | 15 | 21.6 | 14 |
|  | Assam | 60.3 | 16 | 18.4 | 16 |  | Bihar | 63.3 | 16 | 21.5 | 15 |
| India |  | 62.7 |  | 20.4 |  | India |  | 65.4 |  | 23.0 |  |
| Summary Statistics | Mean | 63.10 |  | 20.61 |  | Mean |  | 65.81 |  | 23.36 |  |
|  | SD | 1.57 |  | 1.13 |  | SD |  | 1.65 |  | 1.47 |  |
|  | Mean + SD | 64.67 |  | 21.74 |  | Mean + SD |  | 67.47 |  | 24.83 |  |
|  | Mean - SD | 61.53 |  | 19.49 |  | Mean - SD |  | 64.16 |  | 21.88 |  |

Note: Level of ageing is arranged according to new ageing indicators (RLE15); source: authors' calculations

**Figure 1** and **Table A1** in the appendix indicate that the national trend obscures state-level heterogeneity. The old age threshold of males and females was 62.7 and 65.4 years. To put it another way, the RLE at age 15 differed by around 6 years between Assam and Punjab (Fig. 1 and Table A1 in appendix). For females, there were about 5 years of regional gap in RLE at age 15. The lowest ranked state was Bihar (63.3), while the highest ranked state was Kerala (68.3). We rate the states, with high ranking indicating high ageing and low ranking indicating low ageing. The northern state of Punjab and the southern state of Kerala occupied best rank for males, whereas, Kerala, Punjab, Himachal Pradesh, and Rajasthan occupied best rank for females. Among males, Punjab has the highest RLE (66.9) followed by Kerala (64.5) and lowest values observed in Assam (60.3), Madhya Pradesh (60.7), Uttar Pradesh (61.1). Among females, highest RLE was in Kerala (68.3), followed by Punjab (Punjab), and the lowest in Bihar (63.3) and Orissa (63.8). **Figure 1** also shows that females had better RLE than males in all selected states compared to the males.

**Table 1** compares the ranks of states based on conventional measures of life expectancy at age 60 (LE60) and the new concept of ageing based on the RLE of 15 or fewer years (RLE15). The first rank indicates the highest-ageing state whereas the last rank indicates the lowest-ageing state.

The conventional and prospective measures of ageing ranked the states differently. For example, Karnataka ranked 12th by conventional measures but 10th by the new measure. Bihar ranked 6th on the old metric but 9th by the new. West Bengal dropped from 9th to 12th and Odisha from 11th to 13th, respectively. Among men, Punjab remained ranked 3rd among better-performing states, while Kerala improved to 2nd. Madhya Pradesh declined from 14th to 15th position among the large states using the old and new measures.

While applying the new measure for females, Punjab, Rajasthan, and Assam saw considerable changes in ranks. Punjab went from 4th to 2nd, Rajasthan from 2nd to 4th, and Assam from 15th to 13th. Bihar led major states using the new measure, followed by Odisha and Madhya Pradesh. While measuring from conventional perspective, Assam and Bihar together ranked the lowest followed by Odisha and Madhya Pradesh. Observing gap between males and females, the highest gap of 5 years was found in Rajasthan followed by Kerala (3.8 years), Assam (3.7 years), Himachal Pradesh and Uttar Pradesh (each 3.5 years) (see **Table A1**). While, lowest gap observed in Bihar (0.4 years) followed by Punjab (1.1years) and Odisha (1.2 years).

A rural-urban differential in mortality has long been evident in India. **Figures 2 and 3** compare the urban-rural disparity of RLEs by sex and region for the year 2011. Among the rural males of major states, Punjab (66.2 years), Maharashtra (65.8 years), and Himachal Pradesh (64.3 years) performed better than southern states Kerala (64.2 years) and whereas the worst-performing states were located in the northeastern hilly area Assam (59.7 years), central-eastern belt of Madhya Pradesh (59.9 years) and Uttar Pradesh (59.9 years). Among urban males, the northern state Himachal Pradesh led the way with an RLE=15 at age 69.2, followed by Punjab (67.8) and Haryana (67.6). Uttar Pradesh ranked the least in terms of average RLEs of 15 or fewer after reaching age 63.4, followed by Rajasthan (63.5 years).

Observing females from rural areas, Punjab (68.7) in the north, Rajasthan (68.5) in the west, Kerala (68.4) in the south, Himachal Pradesh (67.9) and Haryana (67.6) in the north performed better. While Bihar (63.1), Maharashtra (63.3), Madhya Pradesh (63.1), Odisha (63.7), and Assam (63.7) were the worst-performing states among rural females (63.8). However, among urban females, Kerala (70.2 years) ranked top, followed by Himachal Pradesh (69.8 years) and Andhra Pradesh (68.7 years). Whereas, Odisha (64.1), followed by Madhya Pradesh (64.3) and Uttar Pradesh (65.1) in the central-eastern region, were the lowest performing elderly females in urban areas. Observing the gap between urban males and females (see **Table A1**), The highest gap between urban males and females was observed in Kerala (5.1), followed by Andhra Pradesh (3.7) and Rajasthan (2.9). On the other hand, for rural area, the male-female gap was highest in Rajasthan (5.8), followed by Haryana (4.4) and Kerala (4.3), and the lowest in Maharashtra (−2.5) followed by Bihar (0.3) and Odisha (1.3). Additionally, rural areas showed a greater female-male gap than urban areas in almost all states except Kerala, Andhra Pradesh, Bihar, and Maharashtra, where the urban-rural gap was slightly larger.

### Comparison of the OADR and POADR by region and sex, India 2011

**Tables 2(a–c)** compares the POADR and OADR in selected major Indian states by place of residence in terms of percentage and absolute number for the period of 2009–2013. It was clear that both the absolute and relative magnitude of ageing greatly increased when POADR was used versus OADR. At the national level, the burden of dependency ratio increased from 8.6% to 10.6% for the overall population. We observed a similar increase for the rural and urban populations. The relative position of the states with respect to dependency ratio also changed when we used the POADR. For example, Kerala had the highest OADR (12.2%) according to the OADR, but ranked 7^th^ and 10.2% when using the POADR (see **Table 2a**).

**Table 2a:** Comparison between the OADR and POADR by overall population (in % and absolute numbers in millions), India 2009–2013.

| Level of ageing | States | OADR x 100 | Old Ranks | Number of old-age dependents (absolute value in millions) | Level of ageing | States | POADR x 100 | New Ranks | Number of old-age dependents (absolute value in millions) |
| --- | --- | --- | --- | --- | --- | --- | --- | --- | --- |
| **High {> (Mean + Standard deviation)}** | Kerala | 12.21 | 1 | 2.78 | **High {> (Mean + Standard deviation)}** | Uttar Pradesh | 12.79 | 1 | 12.60 |
|  | Himachal Pradesh | 10.24 | 2 | 0.47 |  | Orissa | 11.86 | 2 | 2.90 |
| **Moderate (between Mean ± Standard Deviation)** | Maharashtra | 10.01 | 3 | 7.49 |  | Madhya Pradesh | 11.33 | 3 | 4.39 |
|  | Punjab | 9.97 | 4 | 1.87 | **Moderate (between Mean ± Standard Deviation)** | Bihar | 11.24 | 4 | 5.63 |
|  | Tamil Nadu | 9.39 | 5 | 4.73 |  | Andhra Pradesh | 10.93 | 5 | 5.65 |
|  | Orissa | 9.21 | 6 | 2.52 |  | Karnataka | 10.24 | 6 | 3.82 |
|  | Karnataka | 9.04 | 7 | 3.73 |  | Kerala | 10.22 | 7 | 2.18 |
|  | Andhra Pradesh | 8.92 | 8 | 5.12 |  | Maharashtra | 9.95 | 8 | 6.80 |
|  | Madhya Pradesh | 8.37 | 9 | 3.73 |  | West Bengal | 9.87 | 9 | 5.40 |
|  | Uttar Pradesh | 8.31 | 10 | 9.81 |  | Tamil Nadu | 9.86 | 10 | 4.59 |
|  | Haryana | 8.21 | 11 | 1.35 |  | Himachal Pradesh | 9.19 | 11 | 0.38 |
|  | Bihar | 8.2 | 12 | 4.72 |  | Assam | 8.89 | 12 | 1.53 |
|  | West Bengal | 8.13 | 13 | 5.00 | **Low {< (Mean - Standard deviation)}** | Rajasthan | 8.28 | 13 | 2.94 |
|  | Rajasthan | 8.11 | 14 | 3.36 |  | Gujarat | 7.92 | 14 | 2.80 |
| **Low {< (Mean - Standard deviation)}** | Gujarat | 7.71 | 15 | 3.07 |  | Punjab | 7.9 | 15 | 1.32 |
|  | Assam | 6.64 | 16 | 1.30 |  | Haryana | 7.42 | 16 | 1.07 |
| **India** |  | **8.62** |  | **66.43** | **India** |  | **10.57** |  | **71.79** |
| **Summary statistics** | Mean | 8.92 |  |  | **Summary statistics** | Mean | 9.87 |  |  |
|  | SD | 1.25 |  |  |  | SD | 1.49 |  |  |
|  | Mean + SD | 10.16 |  |  |  | Mean + SD | 11.36 |  |  |
|  | Mean - SD | 7.67 |  |  |  | Mean - SD | 8.38 |  |  |

**Table 2b:** Comparison between the OADR and POADR by rural population (in percentage and absolute numbers in millions), rural India 2009–2013.

| Level of ageing | States | OADR x 100 | Old Ranks | Number of old-age dependents (absolute value in millions) | Level of ageing | States | POADR x 100 | New Ranks | Number of old-age dependents (absolute value in millions) |
| --- | --- | --- | --- | --- | --- | --- | --- | --- | --- |
| **High {(> Mean + Standard deviation)}** | Kerala | 12.43 | 1 | 1.47 | **High {(> Mean + Standard deviation)}** | Uttar Pradesh | 13.81 | 1 | 10.28 |
|  | Maharashtra | 12.28 | 2 | 4.83 |  | Maharashtra | 13.42 | 2 | 4.81 |
| **Moderate (between Mean ± Standard Deviation)** | Punjab | 10.86 | 3 | 0.65 |  | Andhra Pradesh | 13.38 | 3 | 4.54 |
|  | Himachal Pradesh | 10.63 | 4 | 0.44 | **Moderate (between Mean ± Standard Deviation)** | Karnataka | 12.64 | 4 | 2.81 |
|  | Karnataka | 10.28 | 5 | 2.55 |  | Orissa | 12.54 | 5 | 2.52 |
|  | Andhra Pradesh | 10.06 | 6 | 3.79 |  | Madhya Pradesh | 12.39 | 6 | 3.36 |
|  | Tamil Nadu | 9.95 | 7 | 2.55 |  | Tamil Nadu | 11.91 | 7 | 2.78 |
|  | Orissa | 9.7 | 8 | 2.18 |  | Bihar | 11.52 | 8 | 5.06 |
|  | Haryana | 8.98 | 9 | 0.94 |  | Kerala | 10.38 | 9 | 1.15 |
|  | Uttar Pradesh | 8.91 | 10 | 7.95 |  | West Bengal | 9.73 | 10 | 3.49 |
|  | Madhya Pradesh | 8.85 | 11 | 2.76 |  | Himachal Pradesh | 9.7 | 11 | 0.36 |
|  | Rajasthan | 8.58 | 12 | 2.59 |  | Punjab | 9.36 | 12 | 0.96 |
|  | Gujarat | 8.49 | 13 | 1.87 |  | Assam | 9.23 | 13 | 1.33 |
|  | Bihar | 8.37 | 14 | 4.21 | **Low {(< (Mean - Standard deviation))}** | Rajasthan | 8.72 | 14 | 2.25 |
| **Low {(< (Mean - Standard deviation))}** | West Bengal | 7.72 | 15 | 3.15 |  | Gujarat | 8.62 | 15 | 1.68 |
|  | Assam | 6.66 | 16 | 1.10 |  | Haryana | 8.4 | 16 | 0.76 |
| **India** |  | **9.21** |  | **47.17** | **India** |  | **11.98** |  | **53.64** |
| **Summary statistics** | Mean | 9.55 |  |  | **Summary statistics** | Mean | 10.98 |  |  |
|  | SD | 1.5 |  |  |  | SD | 1.85 |  |  |
|  | Mean + SD | 11.05 |  |  |  | Mean + SD | 12.83 |  |  |
|  | Mean - SD | 8.05 |  |  |  | Mean - SD | 9.14 |  |  |

**Table 2c:** Comparison between the OADR and POADR by urban population (in percentage and absolute numbers in millions), urban India 2009–2013.

| Level of ageing | States | OADR x 100 | Old Ranks | Number of old-age dependents (absolute value in millions) | Level of ageing | States | POADR x 100 | New Ranks | Number of old-age dependents (absolute value in millions) |
| --- | --- | --- | --- | --- | --- | --- | --- | --- | --- |
| **High {(> Mean + Standard deviation)}** | Kerala | 11.97 | 1 | 1.31 | **High {(> Mean + Standard deviation)}** | Kerala | 9.63 | 1 | 0.98 |
|  | Punjab | 10.22 | 2 | 0.95 |  | West Bengal | 8.74 | 2 | 1.66 |
| **Moderate (between Mean ± Standard Deviation)** | West Bengal | 8.96 | 3 | 1.85 | **Moderate (between Mean ± Standard Deviation)** | Uttar Pradesh | 8.65 | 3 | 2.11 |
|  | Tamil Nadu | 8.81 | 4 | 2.18 |  | Orissa | 8.48 | 4 | 0.37 |
|  | Maharashtra | 7.49 | 5 | 2.66 |  | Madhya Pradesh | 7.89 | 5 | 0.93 |
|  | Madhya Pradesh | 7.25 | 6 | 0.97 |  | Tamil Nadu | 7.82 | 6 | 1.79 |
|  | Karnataka | 7.17 | 7 | 1.19 |  | Andhra Pradesh | 7.42 | 7 | 1.29 |
|  | Bihar | 7.08 | 8 | 0.51 |  | Bihar | 7.35 | 8 | 0.46 |
|  | Himachal Pradesh | 6.97 | 9 | 0.03 |  | Rajasthan | 7.12 | 9 | 0.69 |
|  | Orissa | 6.94 | 10 | 0.34 |  | Maharashtra | 7.09 | 10 | 2.27 |
|  | Haryana | 6.86 | 11 | 0.41 |  | Gujarat | 6.84 | 11 | 1.09 |
|  | Rajasthan | 6.84 | 12 | 0.76 |  | Karnataka | 6.44 | 12 | 0.96 |
|  | Gujarat | 6.74 | 13 | 1.20 |  | Punjab | 6.05 | 13 | 0.39 |
|  | Andhra Pradesh | 6.73 | 14 | 1.33 |  | Assam | 5.96 | 14 | 0.17 |
| **Low {(< (Mean - Standard deviation))}** | Assam | 6.52 | 15 | 0.20 | **Low {(< (Mean - Standard deviation))}** | Haryana | 5.66 | 15 | 0.30 |
|  | Uttar Pradesh | 6.45 | 16 | 1.86 |  | Himachal Pradesh | 4.41 | 16 | 0.02 |
|  | **India** | **7.45** |  | **19.26** |  | **India** | **7.24** |  | **16.66** |
| **Summary statistics** | Mean | 7.69 |  |  | **Summary statistics** | Mean | 7.22 |  |  |
|  | SD | 1.49 |  |  |  | SD | 1.29 |  |  |
|  | Mean + SD | 9.18 |  |  |  | Mean + SD | 8.51 |  |  |
|  | Mean - SD | 6.19 |  |  |  | Mean - SD | 5.93 |  |  |

Source: authors' calculations

According to the OADR, the southern states were the frontrunners in population ageing along with Punjab, Himachal Pradesh, and Maharashtra for the overall population. However, according to the POADR, we found a new set of states with a high dependency on the working-age population. The state-wise distribution of the POADR revealed that Uttar Pradesh had a maximum number of old dependents (12.8%) followed by Orissa (11.9%) and Madhya Pradesh (11.3%). These states had lower ageing based on conventional measures of ageing.

Apart from the above-mentioned examples, we saw ample variation among the states of India in terms of POADR and OADR at the urban-rural level (see **Tables 2b and c**). In all states, we found that the POADR was higher in rural areas than in urban areas. For the rural elderly, it varied between 13.8% in the northern state of Uttar Pradesh followed by the central state of Maharashtra (13.4%; the southern state of Andhra Pradesh (13.4%) ranked the highest. However, we found the lowest values in the northern state of Haryana (8.4%) followed by the western state Gujarat (8.6%) and Rajasthan (8.7%). In urban areas, the situation was slightly better, with the highest percentage of dependent elderly males in Kerala (9.6%) followed by West Bengal (8.7%), Uttar Pradesh (8.6%), and Orissa (8.5%), and the lowest in the northern state of Himachal Pradesh (7.4%) followed by Haryana (5.7%), Assam (5.9%), and Punjab (6.1%). These results clearly indicate that when we used prospective ageing measures, the burden of ageing was higher in the rural than urban population.

We also calculated the absolute number of dependents using both conventional and prospective measures among the major states of India. **Table 2a–c** also compares the absolute number of elderly dependents according to the OADR and POADR by region, India 2011. We found that using prospective measures increased the burden of ageing from OADR 8.6% to POADR 10.6%. There was a huge increase in the percentage of old age to working age individuals of 15–64 years in India, from about 66 million to about 71 million.

According to prospective measures, overall, there were 71 million elderly dependents in India for the year 2011. At the state level, the northern state Uttar Pradesh occupied the highest position with 12.6 million elderlies followed by the central state Maharashtra (6.8 million elderly), southern state Andhra Pradesh (5.64 million elderly), and northern state Bihar (5.63 million elderly). However, the northern states Himachal Pradesh and Haryana occupied the lowest position with 0.3 million and 1.0.6 million elderly dependents, respectively, followed by the northern state Punjab (1.3 million) and eastern state Assam (1.5 million). In rural India, according to the prospective measures, there were about 53.6 million elderly dependents in 2011. At the regional level, it varied between 10.2 million in the northern state Uttar Pradesh and 0.7 million elderly dependents in Haryana. However, in urban India, there were 16.6 million elderly dependents in 2011. At the state level it slightly varied from the rural area. Here, Maharashtra (2.2 million) followed by Uttar Pradesh (2.1 million) had the highest number of elderly dependents in 2011. However, Himachal Pradesh occupied the lowest position with about 19.7 thousand dependents according to prospective measures (see **Table 2a–c**)

## Discussion

Due to the low level of fertility and increasing life expectancy, some states in India have been experiencing ageing faster than other states in recent decades. Researchers and policy makers have recently shown great interest in the issue of ageing and its implications for social and economic aspects of life. However, the majority of previous studies have not taken into account the changing nature of human characteristics with respect to the definition of ageing. The present study estimated the size of the elderly population in India and its regions. This study expands the current literature on the recently developed concept of the characteristic approach of population ageing by Sanderson and Scherbov (2007). To the best of our knowledge, there has been no systematic study that has used the characteristic approach of ageing to examine the detailed heterogeneity of ageing in India.

Our main findings are that both conventional and prospective measures of ageing exhibit wide regional disparity in ageing. However, these indicators measure both the absolute and relative degree of ageing differently for the same population. Considering conventional measures of OADR where the threshold of old age is set to 65 years, we found a higher burden of ageing in India. The limitation of conventional measures is that they consider neither the improvement of life expectancy over time nor the varying degree of life expectancy improvement across regions. Conventional measures of ageing are usually based on the static concept of chronological age since birth, which does not change over time. Many studies have confirmed that continued increases in longevity will ensure that the OADR, which measures the number of elderly people as a share of those who are working age, will rise sharply in most countries over the next 40 years (Muszynska & Rau, 2012). Hence, in a developing country such as India, an old-age dependency rate of 8.6% indicates 66 million elderly dependents in absolute terms, and 9.1% (females) indicates 33 million female dependents in the working age population. However, there has been a consistent increase in life expectancy in India in the past decades along with considerable variation in life expectancies across regions. Therefore, conventional estimates of old-age dependency measures may not reveal the actual size of the elderly. The new prospective measures justify the argument that not only do people get older, they also stay healthy and independent longer in some states in India, whereas for other less-developed states, this argument seems invalid. In other words, increasing life expectancy also means gaining healthy years and not automatically becoming a burden in any population.

Our finding also observed that females have a higher old-age threshold and thus lower POADR in different states, one may argue that this is totally because not considering the health dimension of elderly, however, several studies on disparities in health and life expectancy among fast rising emerging nations such as India and China have shown that when life expectancies increase, disability-free life expectancies increase as well (Thomas, James and Sulaja 2014; Zimmer, Hidajat and Saito 2015). The regional disparity of ageing is evident from our study, in terms of RLE=15, males from Kerala and Punjab and females from Kerala, Punjab and Himachal Pradesh obtained the highest positions in the rankings.

Kerala is undergoing an advanced demographic shift due to declining mortality and fertility rates, and migration plays a vital role in defining the state’s future demographic landscape. In 1989, Kerala became the first Indian state to attain an Infant Mortality Rate (IMR) of less than 25, and the state further decreased the IMR to ten in 2016. However, with an IMR of 34 in 2016, India still has a long way to go before catching up to Kerala. Male and female life expectancies at birth in Kerala were 44.2 and 48.1 years in 1951-60, respectively, and increased to 72.2 and 78.2 years in 2011-15 (Rajan *et al*. 2018). Thus, the state’s age pyramid is rapidly becoming cylindrical to accommodate the ageing population and dwindling youth population. It has a remittance economy, with most of its working population employed overseas, primarily in the Middle East. Moreover, the Middle East oil boom drove massive migration from Kerala to the area. In 2013, there were 2.4 million overseas migrants from Kerala, but that figure fell to 2.2 million in 2016 (Rajan *et al*. 2018). The elderly dependence rate may also have grown as a result of the return of emigrant retirees (James 1994).

According to a recent economic survey of India, with India’s population growth expected to decrease sharply over the next two decades, Punjab and Himachal Pradesh are among the states that will begin transitioning to an ageing society by the 2030s as the number of young people in their population drops due to already low birth rates (GOI 2019). Punjab’s economy is largely agrarian. So, there may be plenty of opportunities for disguised unemployment through work at the family farm, where the marginal productivity of labour is too low to consider. A strong inclination to invest earnings in multiple sectors means that the return on investment may be too large to motivate Punjabis to work. People from Punjab, like Kerala, have a high propensity to work abroad and send remittances; they have mostly travelled to North American nations such as the United States and Canada. This means that Punjab, Himachal Pradesh, and southern India, which are currently leading the demographic change, will also lead the nation’s transition to an ageing society.

We used the POADR as the concept of different ages in different regions, and found that, in contrast to developing countries, applying prospective measures of ageing to high-income countries reduced the size of elderly dependents in both the absolute and relative sense. In India, we found a different scenario in that there were more elderly people in India according to prospective measures compared to the conventional counterpart. This is because life expectancy varies widely across regions of India. The most populous states of India have lower life expectancy, which leads to being old at an early age in those states. The prospective ageing measures not only provided higher estimates of ageing burden in India, but also altered the gender and rural urban differential in ageing.

With this new ageing measure, POADR, we observed a different scenario of ageing in India. This is an interesting measure which takes into account the changes in life expectancies. Increased life expectancy encourages a population to stay in economically productive employment for a significantly longer period of time, which in turn affects policy on retirement age and benefits. According to the POADR, we found a new set of states with a high dependency on the working-age population. The state-wise distribution of the POADR revealed that Uttar Pradesh had a maximum number of old dependents (12.8%) followed by Orissa (11.9%) and Madhya Pradesh (11.3%). These states had lower ageing based on conventional measures of ageing.

The reason might be lies in the fact that Uttar Pradesh being most populous state in India have lower life expectancy compared to southern states. Hence it also has highest elderly population in terms of absolute value. Uttar Pradesh is characterized by extreme variety and disparity in terms of residence, socioeconomic group, gender, and region. The elderly population’s issues exacerbate along these lines; for example, rural elderly inhabitants have more difficulties than urban elderly residents, and elderly women face greater difficulties than male elderly. Moreover, social difficulties such as a prolonged and high rate of widowhood, a lack of social assistance, and an increased reliance on the old population, particularly elderly women, are widespread in this state. Furthermore, elderly health problems deteriorate with age, frequently as a result of neglect, poverty, and social hardship. Poverty and illiteracy deteriorate health and hinder the elder population’s access to health care. Similar argument can be drawn for other least developed states like Orissa and Madhya Pradesh.

This has resulted in an unprecedented burden of ageing in many states, and the issue of ageing must be recognized here. Both the state and the central governments should pay special attention to the older people who are in a less advanced stage and have a poor life expectancy. The present study has also implications in the sense that if we continue to use the traditional dependency ratio, we might risk the true picture of ageing. For instance, RLE in Kerala is very different from Bihar, as LE varies across states in India, thus, it is critical to revisit the traditional method of measuring ageing for various purposes. Additionally, society and family members must be more attentive to the problems and obstacles that the elderly confront.

Our findings contribute interesting knowledge on ageing research in India. Because India is demographically diverse, defining different thresholds of ageing for different regions will change the common discourse on ageing. People living in states with a higher life expectancy at birth may feel that they have aged later than those living in states with a lower life expectancy. This can promote active ageing in states with a higher life expectancy. Moreover, it gives crucial information on policy formulation, as the new concept of ageing can be used at the state level to decide on the retirement age in the public or private sector. Some advanced countries such as Norway, Finland, Spain, Denmark, Greece, Hungary, Italy, Korea, and Turkey have opted to link future increases in pension ages to changes in life expectancy (OECD 2013). More recently, the Netherlands, Portugal, and the Slovak Republic have also linked pension age with life expectancy (OECD 2017). This means that, on average, the retirement period will increase relative to people’s working lives.

To reap the benefits of an experienced and active aging population, governments in various states may review the retirement age. Simultaneously, adequate steps should be made to minimize intergenerational conflict in the labour market. One step in this direction, might be that the government might seek ways to increase the employment capacity of state firms while cutting salary and retirement benefits. If this is realized, two goals may be achieved: dependency reduction and equitable distribution of wealth.

While applying new measures of ageing reduced the size of elderly in demographically advanced states of India, it may also bring about other challenges. Because the unemployment rate is quite high in India among the youth, postponing the retirement age may lead to further unemployment among youth in the government sector.

Our study also highlights the ageing situation in rural India. We found more elderly dependents in rural India compared to urban India. Many rural areas are still remote with poor road and transport access. Income insecurity, lack of adequate access to quality health care, and isolation are more acute in rural elderly than in their urban counterparts. In addition, poorer states such as Uttar Pradesh, Bihar, and Madhya Pradesh have a large absolute number of rural elderlies, which needs special attention. Moreover, rural agriculture has no set retirement age. To reduce reliance in this industry, low-cost loans, suitable technology, and a cost-effective procurement service might be distributed throughout the country. This would maximize the farmer’s return. In the case of land acquisition owing to infrastructure construction, there might be a particular provision for landless workers’ economic rehabilitation.

The present study analyzed state-level variation in ageing, as district-level life expectancies are not available in India. Because intra-state disparity is not negligible in India, there is a scope to expand this study to the district level in the near future. Our analysis also disregarded the socioeconomic disparity across population subgroups.

## Data Availability

All data produced in the present study are available upon reasonable request to the authors

https://censusindia.gov.in/2011-common/censusdata2011.html

https://censusindia.gov.in/vital_statistics/SRS_Statistical_Report.html

https://censusindia.gov.in/vital_statistics/Appendix_SRS_Based_Life_Table.html

## Acknowledgments

Ankita Srivastava, is a recipient of Indian Council of Social Science Research Doctoral Fellowship. Her article is largely an outcome of her doctoral work sponsored by ICSSR. However, the responsibility for the facts stated, opinions expressed and the conclusions drawn is entirely that of the author.

Moreover, part of the research was developed in the Young Scientists Summer Program at the International Institute for Applied Systems Analysis, Laxenburg (Austria) with financial support from the IIASA YSSP FUND 2018. Ms. Srivastava is thankful to YSSP alumni for providing the YSSP Fund 2018.

## Appendix

**Table A1:** Age when RLE=15 among various states of India by place of residence and sex, 2009–2013.

| States | Total Population |  |  | Rural Population |  |  | Urban Population |  |  |
| --- | --- | --- | --- | --- | --- | --- | --- | --- | --- |
|  | Male | Female | Gap | Male | Female | Gap | Male | Female | Gap |
| Andhra Pradesh | 62.6 | 65.7 | 3.0 | 62.0 | 64.8 | 2.8 | 65.0 | 68.7 | 3.7 |
| Assam | 60.3 | 64.1 | 3.7 | 59.7 | 63.8 | 4.1 | 64.6 | 65.8 | 1.2 |
| Bihar | 62.9 | 63.3 | 0.4 | 62.8 | 63.1 | 0.3 | 64.6 | 65.9 | 1.3 |
| Gujarat | 63.6 | 66.6 | 3.1 | 63.1 | 66.8 | 3.7 | 64.3 | 67.0 | 2.7 |
| Haryana | 64.3 | 67.3 | 3.0 | 63.3 | 67.6 | 4.4 | 67.6 | 66.9 | -0.7 |
| Himachal | 64.5 | 68.0 | 3.5 | 64.3 | 67.9 | 3.6 | 69.2 | 69.8 | 0.7 |
| Karnataka | 62.8 | 65.4 | 2.7 | 61.5 | 64.9 | 3.4 | 65.1 | 66.5 | 1.4 |
| Kerala | 64.5 | 68.3 | 3.8 | 64.2 | 68.4 | 4.3 | 65.0 | 70.2 | 5.1 |
| Madhya Pradesh | 60.7 | 63.8 | 3.1 | 59.9 | 63.5 | 3.6 | 64.8 | 64.3 | -0.4 |
| Maharashtra | 64.0 | 66.0 | 2.0 | 65.8 | 63.3 | -2.5 | 65.6 | 66.4 | 0.8 |
| Odisha | 62.6 | 63.8 | 1.2 | 62.4 | 63.7 | 1.3 | 64.0 | 64.1 | 0.2 |
| Punjab | 66.9 | 68.0 | 1.1 | 66.2 | 68.7 | 2.5 | 67.8 | 67.4 | -0.4 |
| Rajasthan | 62.9 | 67.9 | 5.0 | 62.7 | 68.5 | 5.8 | 63.5 | 66.4 | 2.9 |
| Tamil Nadu | 63.3 | 65.3 | 2.0 | 62.4 | 64.6 | 2.2 | 64.7 | 66.4 | 1.7 |
| Uttar Pradesh | 61.1 | 64.6 | 3.5 | 60.6 | 64.5 | 3.9 | 63.4 | 65.1 | 1.6 |
| West Bengal | 62.6 | 65.0 | 2.4 | 61.8 | 64.7 | 2.9 | 64.4 | 65.8 | 1.4 |
| India | 62.7 | 65.4 | 2.6 | 62.0 | 65.0 | 3.0 | 64.8 | 66.4 | 1.6 |

Note: Gap is equal to female minus male age when RLE=15.
Source: authors' calculations

**Table A2:** Expectation of life at birth (e_0_^0^) by sex and residence, India and larger states, 2010–2014.

| India & Larger States | Total |  | Rural |  | Urban |  |
| --- | --- | --- | --- | --- | --- | --- |
|  | Male | Female | Male | Female | Male | Female |
| India | 66.4 | 69.6 | 65.1 | 68.4 | 70.0 | 73.2 |
| Andhra Pradesh | 66.3 | 70.8 | 64.6 | 69.3 | 70.7 | 75.2 |
| Assam | 62.7 | 65.5 | 61.9 | 64.6 | 69.1 | 71.8 |
| Bihar | 67.8 | 68.4 | 67.5 | 68.1 | 70.2 | 71.3 |
| Gujarat | 66.6 | 66.3 | 64.6 | 70.0 | 69.9 | 72.9 |
| Haryana | 66.3 | 71.0 | 65.2 | 70.8 | 68.9 | 72.5 |
| Himachal Pradesh | 69.3 | 74.1 | 68.8 | 73.8 | 75.2 | 77.8 |
| Karnataka | 66.9 | 70.8 | 65.1 | 69.6 | 70.5 | 73.3 |
| Kerala | 72.0 | 77.8 | 71.7 | 78.1 | 72.7 | 77.1 |
| Madhya Pradesh | 62.5 | 66.0 | 61.3 | 65.0 | 67.4 | 70.6 |
| Maharashtra | 69.9 | 73.6 | 68.4 | 72.4 | 71.9 | 75.2 |
| Orissa | 64.7 | 67.1 | 64.1 | 66.5 | 68.7 | 71.0 |
| Punjab | 69.7 | 73.8 | 68.5 | 72.4 | 71.6 | 76.2 |
| Rajasthan | 65.5 | 70.2 | 64.6 | 69.7 | 69.0 | 72.2 |
| Tamil Nadu | 68.6 | 72.7 | 67.2 | 71.3 | 70.5 | 74.5 |
| Uttar Pradesh | 62.9 | 65.4 | 62.1 | 64.7 | 66.8 | 68.9 |
| West Bengal | 68.9 | 71.6 | 68.0 | 70.8 | 71.0 | 73.6 |

Source: SRS ABRIDGED LIFE TABLES- 2010–14
Note: India includes all States/UTs; Andhra Pradesh includes Telangana

